# Effectiveness of the modified WHO labour care guide to detect prolonged and obstructed labour among women admitted at publicly funded facilities in rural Mbarara district, Southwestern Uganda: an ambispective cohort study

**DOI:** 10.1101/2024.09.04.24313073

**Authors:** Mugyenyi R Godfrey, Tumuhimbise Wilson, Atukunda C Esther, Tibaijuka Leevan, Ngonzi Joseph, Kayondo Musa, Kanyesigye Micheal, Musimenta Angella, Yarine T Fajardo, Byamugisha K Josaphat

## Abstract

**Background:** Obstructed labour, a sequel of prolonged labour, remains a significant contributor to maternal and perinatal deaths in low- and middle-income countries.

**Objective:** We evaluated the modified World Health Organization (WHO) Labour Care Guide (LCG) in detecting prolonged and or obstructed labour, and other delivery outcomes compared with a traditional partograph at publicly-funded maternity centers of rural Mbarara district and City, Southwestern Uganda.

**Methods:** Since November 2023, we deployed the LCG for use in monitoring labour by trained healthcare providers across all maternity centers in Mbarara district/City. We systematically randomized a total of six health center IIIs (HCIIIs) out of 11, and all health center IVs (HCIVs), reviewed all their patient labour monitoring records for their first quarter of 2024 (LCG-intervention) and 2023 (partograph-before LCG introduction). Our primary outcome was the proportion of women diagnosed with prolonged and or obstructed labour. Our secondary outcomes included; tool completion, mode of delivery, labour augmentation, stillbirths, maternal deaths, Apgar score, uterine rupture, postpartum haemorrhage (PPH). Data was collected in RedCap and analyzed using STATA version 17. Statistical significance was considered at p<0.05.

**Results:** A total of 2,011 women were registered; 991 (49.3%) monitored using the LCG, and 1,020 (50.7%) using a partograph, 87% (1,741/2011) delivered from HCIVs and 270/2011 (13%) from HCIIIs. Mean maternal age (25.9; SD=5.6) and mean gestation age (39.4; SD=1.8) were similar between the two groups. A total of 120 (12.4%) cases of prolonged/obstructed labour were diagnosed (100 for LCG versus 20 for partograph), with the LCG having six times higher odds to detect/diagnose prolonged/obstructed labour compared to the partograph (aOR=5.94; CI 95% 3.63-9.73, P<0.001). Detection of obstructed labour alone increased to 12-fold with the LCG compared to the partograph (aOR=11.74; CI 95% 3.55-38.74, P<0.001). We also observed increased Caesarean section rates (aOR=6.12; CI 4.32-8.67, P<0.001), augmentation of labour (aOR=3.11; CI 95% 1.81-5.35, P<0.001), and better Apgar Score at 5 minutes (aOR=2.29; CI 95% 1.11-5.77, P=0.025). The tool completion rate was better for LCG compared to (58.5% versus 46.3%), aOR=2.11; CI 95% 1.08-5.44, P<0.001. We observed no differences in stillbirths, maternal deaths, post-partum haemorrhage (PPH) and uterine rupture.

**Conclusions:** Our data shows that LCG diagnosed more cases of prolonged and or obstructed labour compared to the partograph among women delivering at rural publicly funded facilities in Mbarara city/district. We also observed increased C-sections, labour augmentation, and 5-minute Apgar scores. There were no differences in stillbirths, maternal deaths, PPH and uterine rupture. More controlled and powered studies should evaluate the two tools for other delivery outcomes, in different sub-populations.

Trial registration number NCT05979194 *clinical trials.gov*.

**Article Summary:** This manuscript presents novel results from a before-and-after (ambispective cohort study) that utilized retrospective historical data from records of women monitored in labour using an old partograph before introduction of the new modified WHO LCG in South western Uganda. We compared the ability of these labour monitoring tools in detecting cases of prolonged and or obstructed labour and other delivery outcomes at two different times, one year apart. Our data shows that the LCG diagnosed more cases of prolonged and or obstructed labour compared to the partograph, with observed increase in C-section and labour augmentation rates, and no differences in stillbirths, maternal deaths, PPH and uterine rupture. We recommend the LCG as a decision-making tool for use in routine labour in Uganda and similar settings

**Strengths and limitations of this study:**

➢ Our study utilized record reviews which generally represent routine practice and removes the Hawthorne effect where people change/modify or improve their behaviour or practice because they know they are being observed or researched on.
➢ Our retrospective cohort utilized historical partograph records before introduction of LCG in Uganda, while the prospective cohort utilized LCG data at two different times, one year apart, avoiding contamination and observer bias. No known study has reported results comparing clinical outcomes from patients monitored using the old partograph and the new WHO LCG.
➢ Before-and-after designs, also referred to as ambispective cohorts increase statistical power by combining data from multiple sources in a short period of time. Our study presents retrospective partograph data and prospective LCG data.
➢ A small number of records were excluded due to missing critical data on time of onset of labour and time of delivery necessary to robustly define the primary outcome
➢ Due to our preferred study design, we were not able to obtain data on prolonged/obstructed labour detection using the two tools administered to the same mother while monitoring same labour for direct comparison and diagnostic validation.
➢ We were also not powered enough to detect significant differences in maternal deaths, post-partum haemorrhage, uterine rupture and other maternal-foetal outcomes/complications, especially in different maternal demographic or clinical Caesarean section subgroups.

**Implications for implementation and policy:** Our results provide local contextualized data to guide implementation and use of the LCG as an effective decision-making tool in monitoring labor in rural south western Uganda, and similar settings. Health care provider competences in tool use coupled with good implementation strategies in a responsive health care system with good referral networks and LCG champions will improve obstetric outcomes. The results from our study should guide customization of WHO LCG user’s and training manuals to guide roll out of the LCG in Uganda and similar settings to improve intrapartum care for a positive pregnancy and childbirth experience.

## Introduction

Globally Maternal Mortality Ratio (MMR) has declined by 34% over the last decade, from 339 to 223 per 100,000 live births [1]. Although this is commendable, progress towards reaching the Sustainable Development Goals (SDG) 3 target of less than 70 per 100,000 live births by 2030 is still far, given that almost 95% of all these deaths still occur in low- and middle-income countries [2-4]. Uganda has made significant strides towards eradicating maternal and infant mortality rates, with maternal mortality and infant mortality ratio reduction from 336 to 189 per 100,000 live births and from 43 to 34 per 1000 live births respectively [5] Although these efforts are commendable, many women continue to die from preventable pregnancy-related complications, with obstructed labour being a significant contributor to the burden of maternal and perinatal deaths in low resource countries (LRS) [6, 7]. In Uganda, over 8% of all maternal deaths are linked to obstructed labour, with 90% of perinatal mortality following birth asphyxia directly attributed to obstructed labour [8].

To avert these deaths, careful monitoring of labour to timely identify and manage any labour and delivery complications helps to improve birth outcomes [9]. The currently used Friedman’s partograph for labour progress monitoring graphically depicts the dilatation of the cervix, presenting the dilatation of the cervix against time in labour [10, 11]. This labour progress monitoring chart has played a significant role in the past 50 years. However, its utilization and completion rates remain sub-optimal especially in LRS [12-17]. A 2024 systematic review found that the partograph was complex and time consuming, which resulted in low completion rates, and often times non-use among obstetric care workers during labor monitoring in Sub-Saharan Africa, contributing to poor decision making and associated complications during childbirths [18].

After a careful needs assessment, the WHO recommended partograph modification, to fit emerging user and global needs to help reduce the persistently high maternal-and perinatal mortalities and morbidities. The WHO labour care guide (LCG) was then designed to improve assessments for any deviation from normality, and generally monitoring the well-being of women and their babies during labour [19] It is designed to facilitate healthcare providers (HCPs) in quickly identifying deviations from normal labor progress and facilitate timely and appropriate actions to prevent complications. The Ugandan Ministry of Health has now recommended replacing the partograph with the newly recommended LCG [9]. Whereas the LCG has been described as a “next-generation” partograph [20] that incorporates recent effective intrapartum care guidelines, its adaptation to different settings to suite user-needs, and its effect on maternal-foetal outcomes have not been evaluated as a suitable decision-making tool in Uganda.

Between July and December 2023, we conducted stakeholder interviews in rural Southwestern Uganda to identify user needs, customize and refine the new WHO LCG. Preliminary pilot data showed that all 125 HCPs and health care managers interviewed found the modified LCG useful, easy to use, appropriate, comprehensive, appealing and would recommend it to others for labour monitoring [21, 22]. In this study, we assessed the effect of the modified LCG in detecting prolonged and or obstructed labour, and other labour/delivery outcomes compared to the traditional partograph at public basic and comprehensive emergency obstetric and newborn care (B/CeMONC) facilities of rural Mbarara district and City, Southwestern Uganda.

## Methods

### Study Design

This was a retrospective review of labour and delivery records for all mothers delivering at selected public health facilities offering basic and comprehensive emergency obstetric care in Mbarara district and Mbarara City. We compared delivery outcomes from an ambispective (before - and -after) study that comprised of a prospective cohort of labour records of women monitored using the modified LCG (intervention arm) and a historical cohort of women monitored using the partograph before the introduction of the LCG (comparison arm).

### Study setting

We included all public health centres offering basic and comprehensive emergency obstetric and newborn care in Mbarara district and city. These include all 11 public health centre IIIs (HCIIIs) located in each subcounty, two health centre IVs (HCIVs) (Bwizibwera, Mbarara City Council). The district is served with a total of 253 Hhealth care providers (HCPs) who provide obstetric healthcare, with a large concentration at Mbarara Regional Referral Hospital (MRRH). Each HCIV has 2–3 medical officers and 10 midwives on average [23] with HCIIIs having 3–5 midwives each.

This study collected data from patients’ charts in the labour suites and postnatal wards of six HCIIIs, including Bubaare, Rubaya, Rubindi in Mbarara district; Biharwe, Nyakayojo, and Kyarwabuganda distributed Mbarara city. Before the start of the study, all mothers in labour were ideally monitored using a partogram and the fetal heart rate measured manually per clinician judgement using the Pinard stethoscope. After a normal (uncomplicated) vaginal delivery, the mothers from these facilities with their babies are admitted to the postnatal wards for 24 hours, with daily ward rounds conducted by skilled birth attendants. Those who deliver by caesarean section remain admitted for 3–5□days though mothers and babies with complications are admitted for more days.

Mbarara district is located approximately 270 km southwest of the capital, Kampala, with a population of about 250□000 people distributed through two recent administrative units of Mbarara city and Mbarara district. Uganda’s public health system is organised into seven tiers with national and regional referral hospitals, general district hospitals and four levels of community health centres. Staffing and available services vary across the four levels: HCIII offers basic emergency obstetric care (carry out prenatal care and conduct vaginal deliveries), whereas HCI and HCII serve as low-resource primary healthcare units. HCIVs and hospitals conduct normal vaginal and caesarean deliveries (offer comprehensive emergency obstetric care), and have ambulances and blood transfusion services [24]. Private providers operate in parallel to the public health system to provide maternal healthcare. The district has one publicly funded referral hospital-MRRH, which doubles as a teaching hospital for Mbarara University of Science and Technology (MUST), plus two HCIVs of Bwizibwera and Mbarara city. The prenatal care attendance of ≥4 visits is still at 58%, and maternity services, including delivery, are largely provided free of charge.

### Study procedure

From November 2023 to date, we deployed the recently adapted LCG [21, 22] for use in routine labour monitoring by HCPs at all public health facilities offering basic and comprehensive emergency obstetric care in Mbarara district and Mbarara City following training. These included all eleven HCIIIs (Bubaare, Bukiro, Kagongi, Kashare, Rubaya and Rubindi in Mbarara district and Biharwe, Kakoba, Nyakayojo, Nyamitanga and Kyarwabuganda in Mbarara City, plus the two HCIVs (Bwizibwera and Mbarara City Council), and MRRH. Labour monitoring was done by skilled HCPs who were trained for proficiency in the use of labour care guide. We systematically randomized a total of six HCIIIs-half of the total (three in Mbarara City-Biharwe, Kyarwabuganda, Nyakayojo and three in Mbarara district-Rubindi, Rubaya, Bubaare). We included all the two HCIVs. These rural facilities generally have a stable and or firmly fixed community and population of HCPs offering prenatal, labour and delivery services to their catchment areas. MRRH was excluded in this data review because it participated centrally in the adoption, refining, training and pilot testing of the original WHO LCG to the final Ugandan prototype for evaluation. It also has a high turnover of HCPs, being a teaching hospital for MUST, with a dynamic or unstable population of HCPs including obstetrics and gynaecology residents and midwifery students.

Following the launch of the Uganda Essential Maternal and Newborn (EMNC) guidelines 2022, Ministry of Health conducted country wide dissemination of these new guidelines targeting all HCPs at all public basic and comprehensive emergency maternal and newborn centres. Experienced LCG trainers, delivered the LCG modules to all HCPs. Following modification and refining [21, 22] the tool was provided to all public facilities in Mbarara for use during routine labour monitoring. Additional training for each site on LCG use was done by our research team in October 2023 before final tool deployment. The partographs in Mbarara were recalled and replaced with the LCG to record labour progress and delivery outcomes.

Research assistants who were trained in human subjects’ protection (HSP), labour care guide use, study protocols and data collection tools retrieved all labour and delivery records from the records sections of the eligible health facilities to the data collection centre. We pilot tested and refined the data extraction form on 50 independent delivery records (25 for women monitored using the partograph and 25 for using the modified LCG). The refined form was uploaded in REDcap for data collection. Study variables of interest were extracted from all labour records of patients in same facilities monitored using the modified LCG for the first quarter of 2024 (January-March 2024 for LCG-intervention arm) and first quarter of 2023 (January-March 2023 for partograph-Comparison arm) regardless of parity, maternal age, gestation age and referral status. The year 2023 is a period prior to introduction of the LCG in Uganda.

We screened all labour monitoring tools to exclude intrauterine fetal deaths, multiple gestation and breech presentations. We also excluded those forms with missing time of onset of labour and time of delivery.

### Study outcomes

The primary outcome of interest was women diagnosed with prolonged and or obstructed labour. In this study, obstructed labour was defined as failure of labour to progress despite presence of adequate uterine contractions, due to mechanical reasons usually a mismatch between the size of the baby and the pelvis. We defined prolonged labour as labour crossing the action line on the partograph (for women monitored with partograph) a labour lasting more than a specified centimetre cervical dilation “time lag” in the alert column of section five of the labour care guide. The labour monitoring section of each tool was assessed and categorized as normal labour, prolonged labour, obstructed labour or a missed diagnosis of prolonged and/or obstructed labour. Our secondary outcomes included tool completion rate, mode of delivery, proportion of women who received labour augmentation, stillbirths, maternal deaths, Apgar score, uterine rupture and PPH.

All labour monitoring tools (LCG or partograph) were subjected to a completeness check to assess the level of filling labour observations. A labour monitoring tool was regarded to be complete or filled to standard when it fulfilled all the three major criteria plus any two minor criteria, while an incompletely filled tool missed recording any of the three major criteria. All data for each labour/delivery record was double-checked with a different research assistant to minimize errors.

A labour monitoring tool was regarded to be completely filled or filled to standard if it fulfilled the following major and minor criteria. The major criteria included recording; 1) Fetal Heart Rate (FHR) continuously every 30 minutes throughout active labour on the partograph and the LCG; 2) the number or frequency and duration of uterine contractions continuously every 30 minutes throughout active labour on the partograph (shading contractions according to frequency/number and duration) and the LCG; 3) at least two columns completely filled throughout section 1-6 of the LCG or four sections of the partograph including record of fetal wellbeing (FHR, character of liquor, and moulding), labour progress (cervical dilatation, descent and uterine contractions), and record of maternal wellbeing (blood pressure, pulse rate). All labour observations should be recorded at each assessment including at least during admission in active labour (baseline) and at the onset of second stage of labour.

The minor criteria included recording; 1) Prevention of Mother to Child Transmission (PMTCT) code for HIV (space for recording HIV test results/code is provided on both the partograph and LCG); 2) Duration of active labour (there is space for recording duration of active labour or at least calculate it); 3) Amount of blood loss; 4) Post-delivery blood pressure; 5) Newborn temperature. There is space provided to record these on both the LCG and the partograph. Post-delivery blood pressure is an early predictor of post-partum haemorrhage, the leading cause of maternal mortality.

### Sample Size determination

We used an online formula for independent cohort studies [25-27]. The goal was to detect at least a 10% difference in prolonged/obstructed labor diagnosed between the two groups of women monitored using the LCG and the traditional partograph, with a power of 90% and a two-sided type I error of 5%. The assumed incidence of prolonged labor with the partograph was 8%[8]. Using these parameters, the study required a minimum of 520 participants in each group to achieve the desired power and detect the differences in prolonged/obstructed labor between the two groups.

### Statistical Analysis

We followed Consolidated Standards for Reporting Trials (CONSORT) guidelines 2010[28] to conduct and report the findings. We summarized and described the socio-demographic and other health-related data between the two groups: the prospective LCG cohort and the historical partograph. The primary outcome was the proportion of women diagnosed with prolonged/and obstructed labor. Secondary outcomes included labor augmentation, mode of delivery, still births, maternal death, PPH, Apgar Score, uterine rupture and tool completion.

We considered prolonged/obstructed labour as a dichotomous outcome and described the performance of the LCG compared with the partogram-monitored historical cohort. We fitted a multivariable logistic regression model, with study group/arm as the predictor of interest, and controlled for maternal age, parity, HIV status, referral status and health facility level at the time of enrolment, due to not only their strong association with prolonged labour but also differences at enrolment between the two groups. Although not designed to detect a difference, also explored additional secondary outcomes as listed above.

#### Interaction term analysis

To understand the combined effect of some clinical and demographic characteristics on the primary outcome variable, we performed interaction term analysis to identifying the effect of each exposure variable on detecting prolonged/obstructed labour in women monitored using the LCG and the partograph. Data analysis was conducted using STATA version 17. (Statacorp, College Station, Texas, USA). Statistical significance was considered at p ≤ 0.05.

### Ethics Statement

This study was approved by the Institutional Ethics Review Committee of Mbarara University of Science and Technology (MUST-2023-808) and the National Council for Science and Technology in Uganda (UNCST–HS2864ES). Additionally, the study obtained permission from the Director curative services of the ministry of health, administrative site clearance from Mbarara City and Mbarara District Health Officers. Before review of patient labour records, we obtained a waiver of consent from Mbarara University Research Ethics Committee. The study was conducted in compliance with the ethical standards of responsible conduct of research on human subjects, as per the Helsinki declaration.

## Results

Of the 2,092 records of women who were monitored for delivery from January to April 2023 (partograph) and January to April 2,024 (LCG), 2,055 records were screened for eligibility for data abstraction (Figure 1). Thirty-seven (37) records were ineligible because mothers presented with intrauterine fetal death (04), breech presentation (15) and multiple gestation (18) at onset of labour. Forty-four (44) records were excluded because of missing labour and delivery data; [including time of onset of labour (23) and time of delivery (21)], which are critical to define the primary outcome. A total of 2,011 records were included in the final analysis, with 991 (49.3%) monitored using the LCG, and 1,020 (50.7%) using the partograph. Clinical and demographic characteristics including health facility level, maternal age, gravidity, parity, gestational age and HIV status were similar in both groups, except mothers’ referral status and cadre of HCP conducting the delivery (Table 1). Up to 1,739 (86.5%) women delivered from HCIVs and 272 (13.5%) from HCIIIs. The mean maternal age (standard deviation [SD]) was 25.9 (5.6) years, with mean gestation age (SD) of 39.4 (1.8) weeks (Table 1).

A total of 120 (12.4%) cases of prolonged and/or obstructed labour were diagnosed, with 100 (10.4%) cases detected using the LCG versus 20 (2.0%) cased detected using a partograph (Table 2). This translates to six times high odds of LCG’s ability to diagnose obstructed and or prolonged labour compared to the partograph (aOR=5.94; 95%CI 3.63-9.73 P<0.001) when we adjusted for parity, maternal age, cadre conducting delivery, referral status and HIV status. The LCG was also four times more likely to diagnose prolonged labour alone compared to the partograph (aOR=3.75; CI 95% 2.15-6.54, P<0.001). Detection of obstructed labour alone increased to 12-fold when compared between LCG and partograph (aOR=11.74; CI 95% 3.55-38.74, P<0.001). The incidences of labour augmentation (aOR=3.11; CI 95% 1.81-5.35, P<0.001), and Caesarean Section delivery at HCIVs (aOR=6.12; CI 95% 4.32-8.67, P<0.001) also increased by three and 6-fold respectively for women monitored by LCG compared to the partograph. We also noted more babies (94% versus 90%) reported in the LCG group whose Apgar score was greater than seven compared to the babies whose mothers were monitored using partograph group (aOR=2.29; CI 95%1.11-5.77, P=0.025). Lastly, also still low, we observed a higher completion rate in filling out an LCG compared to the partograph (aOR=2.11; CI 95% 1.08-5.44), P=0.001). The LCG showed two times more likely to be completed or filled to standard (580/991, 58.5%) compared to the partograph (472/1,020, 46.3%; aOR=2.11; CI 95% 1.08-5.44, P=0.001).

We analysed for other delivery outcomes. Although not powered to detect the differences, we observed no differences in stillbirths (aOR=0.36; CI 95% 0.06-1.79, P=0.074), maternal deaths (aOR=2.46; CI 95% 0.25-23.92, P=0.437), postpartum haemorrhage (aOR=2.1; CI 95% 0.7-5.90, P=0.177), Apgar score at one minute (aOR=0.75; CI 95% 0.41-1.40, P=0.370) and uterine rupture (aOR=0.54; CI 95%. 0.17-2.82, P=0.093). While we did not power our study to estimate differences within sub-groups, our data shows slight differences in prolonged/obstructed labour incidences across specific demographic and clinical characteristic sub-groups of mothers monitored with LCG versus partograph such as HIV status, parity, referral status, gestation age, facility level and cadre monitoring and or conducting the delivery (Table 3). However, the overall differential odds of detecting obstructed and or prolonged labour remains higher in the LCG compared to the partograph group.

Lastly, the stratified analysis to assess for differences in our primary outcome within the predictive baseline demographic and clinical sub-groups, found that none of the sub-group-by-labour monitoring tool interaction terms was significant (Table 3).

## Discussion

In this ambispective study that compared a prospective cohort of labour records of women monitored using the new modified WHO LCG (intervention) and a historical cohort of women monitored using the traditional partograph (comparison group), we demonstrated that the LCG was better in detecting prolonged and or obstructed labour. The LCG detected or diagnosed 10.4% cases of prolonged and or obstructed labour compared to the partograph, which detected only 2.0% of the cases. In fact, the LCG was found to detect or diagnose six times more cases of obstructed and or prolonged labour combined compared to the partograph, while detecting 12 times more cases of obstructed labour alone. Interestingly, we also found increased proportions of women that received labour augmentation and those that delivered by Caesarean section at HCIVs in the LCG category compared to women monitored by the partograph. In fact, the odds of labour augmentation and Caesarean sections increased by three and 6-fold in the LCG cohort when compared to the partograph cohort respectively. There were also more babies with Apgar score greater than seven at 5 minutes (94% versus 90%) in the LCG group compared to the women monitored using the partograph, with LCG completion rate better compared to that for the partograph. However, we observed no significant differences in still births, maternal deaths, PPH, obstetric shock index and uterine rupture, and none of the sub-group-by-labour monitoring tool interaction terms was significant.

In 2018, WHO provided guidance on care of mothers in labour in the context of a positive child birth experience [29, 30], with the partograph imminently undergoing massive revision to enable care basing on the emerging evidence and global priorities [31-33]. Our data provides us with local contextualized evidence required to further understand the use and effectiveness of the labour care guide in Uganda and similar settings. In this ambispective cohort, we used a customized LCG, a recent modification of the WHO labour care guide to align with MOH’s reproductive health care programs, printed back to back on an A4 as a one stop quick reference document to aid clinical decisions, team work, accountability and interaction among HCPs, women and others on the care team like the birth companion and family members [21, 22]. The observed significant improvement in recognising and timely detecting/diagnosing prolonged labour could be attributed to the LCG’s embedded “alert column criteria” that seems to provide HCPs a “cue to action”, activating them to critically analyse each recorded labour observation for any deviations from normal for timely interventions before obstructed labour sets in [12, 34]. The partograph on the other end has been seen to largely depend on the 4-hourly cervical dilatation assessment and its plot reaching the alert and action line [10, 35-37].

The ability of the LCG to detect six times more cases of prolonged and obstructed labour combined, detecting 12 times more cases of obstructed labour alone, compared to the women monitored using partograph is of great importance. This high detection probably offered opportunity for timely interventions such as labour augmentation and Caesarean section to avert impending labour complications, in the LCG group compared to the partograph. Our observed increased incidences of labour augmentation and delivery by Caesarean section therefore, could have been due to these dynamics aimed at seeking timely interventions, with more babies reportedly born with Apgar score greater than at 5 minutes, and no reported uterine rupture in the LCG group compared to the partograph. These dynamics could indeed make the LCG a potentially feasible and acceptable tool for labour monitoring in varying clinical settings such as Uganda [38].

Whereas a stepped-wedge cluster-randomized trial to introduce the LCG into routine care in India found a lower Caesarean section rate at 39.7% in the LCG compared to 45.2% in the comparison group, it is important to note that aside from segregating and selectively reporting the Caesarean sections in Robson’s category 1, Vogel and colleagues also conducted their study in tertiary hospitals. This presents significant differences with rural community health facilities, with limited capacity and expertise in our study. It is therefore possible that in this tertiary hospital setting, there were more advanced skilled birth attendants like OBGYN residents compared to our setting where the majority were delivered by enrolled certificate-level midwives routinely deployed at rural community health facilities. Additionally, Robson’s group one category includes nulliparous women with cephalic, singleton pregnancies great than 37 weeks of gestation with spontaneous labour [39, 40]. Our study also included records of all women who went into active phase of labour with live foetuses who were monitored at a typical basic and comprehensive emergency obstetric and newborn care centres in Uganda, irrespective of parity, maternal age, gestational age, prior obstetric history, labour onset, fetal presentation, HIV status, cadre conducting the delivery or whether labour was induced or spontaneous. Only women with IUFD and multiple pregnancy were excluded in our study. We also think that in the stepped-wedge trial, HCPs were under close observation and possibly not blinded to the study’s hypothesis, factors that could have modified their behaviour simply because they knew they were being observed or knew what was being assessed (Hawthorne effect) [41]. In our study all HCPs were trained to ensure LCG use and proficiency, and then left to use the tool routinely to monitor labour without further observation. We, however, observed no differences in still births, maternal deaths, PPH, obstetric shock index, Apgar score at one minute and uterine rupture, in line with Vogel and colleagues’ study [39, 40].

The observed higher completion rate in filling out the LCG compared to the partograph could also have provided a good edge to diagnose prolonged labour and other delivery indices, facilitating timely action and interventions. Comparatively, a study of 527 parturients at a regional referral hospital in Southwestern Uganda found an abysmal 4.2% partograph completion rate [16]. Noteworthy, our results report higher tool completion rates of 58.5% (LCG) vs 46.3% (partograph) filled to a set standard for all eligible records of women monitored for labour at lower-level, less busy community health facilities, with a stricter and recommended criterion for labour monitoring [19]. Whereas Lugobe et al considered completeness to standard as a record of 1) cervical dilatation recorded at least once every 4 hours; 2) fetal heart rate recorded at least once every hour; and 3) uterine contractions recorded at least once every hour [16, 42, 43] on the partograph, our study developed and used three major and five minor criteria for a labour monitoring tool to be considered as standard. Our major criteria included 1) Fetal Heart Rate recorded continuously every 30 minutes throughout active labour on the partograph and LCG, 2) the number or frequency and duration of uterine contractions recorded continuously every 30 minutes throughout active labour on the partograph and LCG and 3) at least two columns completely filled throughout section 1-6 of the LCG or 4 sections of the partograph. We also used the five minor criteria including a) record of prevention of Mother to Child Transmission (PMTCT) code for HIV, b) duration of active labour, c) amount of blood loss, d) post-delivery blood pressure and f) newborn temperature. These criteria cut across uniformly in all the two labour monitoring tools, and space is provided for recording all these important labour and delivery observations on both tools as part of routine observations consistent with MOH programs to improving pregnancy outcomes.

A realist review examines what it is about a particular practice or intervention that works/ is useful or does not work, who it works for, and in which settings it works best, ranging from support to health systems, acceptability by health workers, effective referral mechanisms, human resources and health provider competence [44]. The higher completion rate of LCG was expected due to a new enthusiasm with the new tool, HCP empowerment through adequate training and mentorship at our study sites. The completeness rates for the LCG were, however, lower than expected, attributed to poor attitude and documentation practices reported by other scholars in similar settings [16, 42, 43]. This failing HCP attitude and practices could be improved through targeted incentives, ongoing support supervision and mentorships [21, 22]. Whereas studies using realist review methods have shown that directly incentivising individual HCPs to use the partograph did not translate into sustained practice, clinical stewardship, ongoing support supervision, empowerment, intrinsic and acquired professional skills/competence and the organisational setting in the wider context of improving obstetric care and referral were key to full and sustained acceptability and utilization of monitoring tools [45]. To improve completion rates and so improving labour and delivery outcomes while using the new LCG, particular attention should therefore, be directed to critical end-user perspectives, and use these perspectives to design appropriate implementation strategies at individual (training, attitudes), health system and intervention (tool itself) levels to attain improved pregnancy outcomes.

Our study has several strengths and paints a typical picture of use of labour monitoring tools in busy community facilities offering routine basic and or comprehensive obstetric care, subject to considerable publicly funded challenges in rural Southwestern Uganda. The retrospective cohort utilized the old partograph data before any introduction of LCG in Uganda, while the prospective cohort utilized the customized LCG data following training, LCG deployment and partograph recall across all facilities in the study site at two different times, one year apart, avoiding contamination and observer bias. Our ambispective cohort evaluated delivery outcomes of an intervention in a fairly fixed and similar population of HCPs at HCIII and HCIVs where more than 50% of deliveries in Uganda occur [46], making our results generalizable in similar settings. Our study therefore provides novel, local context and first-hand findings and evaluation between the customized “new generation partograph” described as simpler, easier to use, comprehensive, and the old traditional partograph, presenting effectiveness data to support use of the LCG against the standard (partograph) as a decision-making tool in monitoring labour and detecting prolonged/obstructed labour.

Record reviews generally represent routine practice and removes the Hawthorne effect where people change/modify or improve their behaviour or practice because they know they are being observed or researched on. Additionally, ambispective cohorts increase statistical power by combining data from multiple sources in a short period of time-retrospective partograph data and prospective LCG data. We included all data for a whole quarter, enabling us to robustly and timely evaluate effectiveness of the LCG compared to the partograph, while ensuring statistical validity for meaningful conclusions.

In our study, a small number of records were excluded due to missing data (2%, 44/2092) on time of onset of labour and time of delivery necessary to define prolonged labour. A small number (1.8%, 37/2092) was also excluded due to presence of intrauterine fetal deaths, multiple gestation and breech presentations. On the other hand, we included every data regardless of parity, maternal age, gestational age, prior obstetric history, labour onset, fetal presentation, HIV status, cadre conducting the delivery, assessed all Caesarean deliveries regardless of Robson class, or whether labour was induced or spontaneous, thus improving the potential for generalizability of our findings. Our research team also developed and used a standard criterion for describing and evaluating tool completeness check based on standard or routine and pragmatic labour and delivery measurements and outcomes, making our data reliable.

Our study also had a few limitations. Due to our preferred study design, we were not able to obtain data on prolonged/obstructed labour detection from the two tools administered to the same mother while monitoring same labour for direct comparison and diagnostic validation. However, we think our effectiveness data is very informative to the MOH’s proposed policy change. We were also not powered to detect significant differences in maternal deaths, post-partum haemorrhage, uterine rupture and other maternal-foetal outcomes/complications, especially in different maternal demographic or clinical Caesarean section subgroups.

## Conclusion and Recommendations

Our data shows that LCG had six times more chances of detecting/diagnosing obstructed and or prolonged labour combined compared to the partograph. Our data also shows that the LCG had 12 times more chances of diagnosing a case of obstructed labour alone when compared to the partograph among women delivering at rural public health facilities in Uganda, Southwestern Uganda. Although we were not powered to detect the differences, our data showed increased interventions such as C-Sections at CeMONC facilities, and labour augmentation, and better Apgar score at 5 minutes. The tool completion rate was better in the LCG group compared to the partograph group. We found no differences in still births, maternal deaths, PPH and uterine rupture. More controlled and well powered studies should evaluate the two labour monitoring tools for tool completion other labour and delivery outcomes, and in different maternal and HCP sub-populations.

## Supporting information

Table 3: Maternal characteristic sub-groups by LCG versus partograph with prolonged/ obstructed labor distribution

Table 2: Primary and secondary outcomes compared by LCG versus Partograph

Table 1: Socio-demographic and obstetric characteristics of participant

## Data Availability

All data produced in the present study are available upon reasonable request to the authors

## Author Contributions

**Conceptualization:** Godfrey R. Mugyenyi, Josaphat K. Byamugisha, Esther C. Atukunda, Yarine T. Fajardo. **Data curation:** Godfrey R. Mugyenyi, Wilson Tumuhimbise, Leevan Tibaijuka, Kanyesigye Micheal **Formal analysis:** Godfrey R. Mugyenyi, Leevan Tibaijuka, Esther C. Atukunda. **Methodology:** Godfrey R. Mugyenyi, Josaphat K. Byamugisha, Esther C. Atukunda, Yarine T. Fajardo. **Project administration:** Godfrey R. Mugyenyi. **Resources:** Godfrey R. Mugyenyi, Esther C. Atukunda. **Supervision:** Josaphat K. Byamugisha, Yarine T. Fajardo. **Writing – original draft:** Godfrey R. Mugyenyi. **Writing – review & editing:** Josaphat K. Byamugisha, Wilson Tumuhimbise, Kanyesigye Micheal, Leevan Tibaijuka, Ngonzi Joseph, Kayondo Musa, Musimenta Angella, Esther C. Atukunda, Yarine T. Fajardo.

## Availability of data and materials

De-identified REDCap CSV/Microsoft Excel (raw data) files have been provided alongside this submission. Appropriate data sets will be available on reasonable request from the authors. Data sets will be deposited in a repository for future reference.

## Acknowledgments

NIH funded Mbarara University of Science and Technology support-Moms Project Uganda, (Grant number1R01HD111692-01) for funding the field visits and data collection, Micheal Kanyesigye for developing and maintaining the REDcap database. Evelyne Kahumuro, Prossy Tusiime, Ddamba Frank, Bajja Uthman, Sandra Chepkwemoi

## Trial Registration

This trial registration was registered with *<u>clinical trials</u>.<u>gov</u>* number NCT05979194 on 2023-08-07, and the protocol was published by BMJ open, as 10.1136/bmjopen-2023-079216 on 15 April 2024 [22]

