## Supplementary material for "Effectiveness of the modified WHO labour care guide to detect prolonged and obstructed labour among women admitted at publicly funded facilities in rural Mbarara district, Southwestern Uganda: an ambispective cohort study": Table 3: Maternal characteristic sub-groups by LCG versus partograph with prolonged/ obstructed labor distribution

| **Characteristic** | | **LCG (N=991)** | **Partograph (N=1,020)** | **OR (95% C.I)** | **p-value** | **p-value for Interaction Term** |
| --- | --- | --- | --- | --- | --- | --- |
| HIV status | |  |  |  |  |  |
|  | Negative | 82/814 (10.1%) | 16/804(2.0%) | 5.52(3.20-9.52) | <0.001* | 0.839 |
|  | Positive | 6/70 (8.6%) | 1/75 (1.3%) | 1.26(0.13-11.48) | 0.942 |  |
|  | Unknown | 12/107 (11.2%) | 3/141 (2.1%) | 4.07(0.31-3.72) | 0.915 |  |
| Parity | |  |  |  |  |  |
|  | Primipara | 43/448 (9.6%) | 11/391 (2.8%) | 3.36(1.86-7.23) | <0.001* | 0.493 |
|  | 2-4 | 19/297 (6.4%) | 3/266 (1.1%) | 5.23(1.76-10.50) | <0.001* |  |
|  | >5 | 3/27 (11.1%) | 2/33 (6.1%) | 1.63(0.40-6.65) | 0.145 |  |
|  | Unknown | 35/219 (16.0%) | 4/330 (1.2%) | 4.23(1.21-14.74) | 0.024* |  |
| Referred in | |  |  |  |  |  |
|  | No | 81/937 (8.6%) | 18/1011 (1.8%) | 5.22(3.12-8.77) | <0.001* | 0.257 |
|  | Yes | 19/54(35.2%) | 2/9 (22.2%) | 15.76(3.06-21.99) | 0.001* |  |
| Gestational age | | | | | |  |
|  | 28-36 weeks | 3/35 (8.6%) | 0/27 (0) | - | - |  |
|  | 37-42 | 44/484 (9.1%) | 9/473 (1.9%) | 5.23(3.07-7.45) | <0.001* | 0.983 |
|  | >42 | 53/472 (11.2%) | 11/520 (2.1%) | 5.78(3.20-9.76) | <0.001* |  |
| Facility Level | | | | | |  |
|  | Health Centre IV | 96/845 (11.4%) | 18/894 (2.0%) | 6.22(3.73-10.39) | <0.001* | 0.169 |
|  | Health Centre III | 4/146 (2.7%) | 2/126 (1.6%) | 1.78(0.18-3.42) | 0.747 |  |
| Cadre conducting the delivery | | | | | |  |
|  | Certificate Midwife | 27/621(4.3%) | 14/794 (1.8%) | 2.53(1.32-4.87) | <0.001* | 0.544 |
|  | Others | 73/370(19.7%) | 6/226(2.7%) | 6.04(4.91-8.36) | 0.007 |  |
