## Supplementary material for "Effectiveness of the modified WHO labour care guide to detect prolonged and obstructed labour among women admitted at publicly funded facilities in rural Mbarara district, Southwestern Uganda: an ambispective cohort study": Table 2: Primary and secondary outcomes compared by LCG versus Partograph

|  |  | **LCG (N=991)** | **Partograph (N=1,020)** | **Unadjusted analysis** | | **Adjusted analysis** | |
| --- | --- | --- | --- | --- | --- | --- | --- |
| **Outcome** | |  |  | ***c* OR (95% C.I)** | **p-value** | ***a* OR (95% C.I)** | **p-value** |
| **Primary outcome** | |  |  |  |  |  |  |
|  | Combined obstructed and prolonged labour | 100 (10.4%) | 20 (2.0%) | 5.61(3.44-9.15) | <0.001* | 5.94(3.63-9.73) | <0.001* |
|  | Prolonged labour alone | 64 (6.6%) | 17 (1.7%) | 4.07(2.37-7.01) | <0.001* | 3.75(2.15-6.54) | <0.001* |
|  | Obstructed labour alone | 36 (3.7%) | 3 (0.3%) | 12.78(3.92-41.63) | <0.001* | 11.74(3.55-38.74) | <0.001* |
| **Secondary outcomes** | | | | | | | |
|  | Labor augmentation | 52 (5.3%) | 17 (1.7%) | 3.27(1.90-5.61) | <0.001* | 3.11(1.81-5.35) | <0.001* |
|  | Caesarean section delivery | 179 (18.1%) | 54 (5.3%) | 3.94(2.87-5.42) | <0.001* | 6.12(4.32-8.67) | <0.001* |
|  | Still births | 22(2.2%) | 25(2.5%) | 0.79(0.48-1.85) | 0.200 | 0.36(0.06-1.79) | 0.074 |
|  | Maternal death | 3 (0.3%) | 1 (0.1%) | 3.09(0.32-29.70) | 0.328 | 2.46(0.25-23.92) | 0.437 |
|  | PPH | 15 (1.5%) | 5 (0.5%) | 3.1(1.1-8.6) | 0.028* | 2.1(0.7-5.9) | 0.177 |
|  | Mean Obstetric Shock index (SD) | 0.751(0.11) | 0.747(0.10) | N/A | N/A | N/A | N/A |
|  | Normal Apgar Score >7 at 1 minute | 916 (92.4%) | 925(90.7%) | 1.28(0.04-2.52) | 0.277 | 0.75(0.41-1.40) | 0.370 |
|  | Normal Apgar Score>7 at 5 minutes | 935 (94.3%) | 922(90.4%) | 1.48(1.17-3.68) | 0.003* | 2.29(1.11-5.77) | 0.025* |
|  | Uterine rupture | 0(0) | 2(0.2%) | 0.32(0.04-1.52) | 0.160 | 0.54(0.17-2.82) | 0.093 |
|  | Tool completion rate | 580(58.5%) | 472(46.3%) | 1.55(1.11-3.47) | 0.003* | 2.11(1.08-5.44) | 0.001* |
| *LCG: Labor care guide, c OR: crude Odds Ratio, aOR: adjusted Odds Ratio, C.I: Confidence Interval, PPH: postpartum haemorrhage, SD: Standard Deviation* | | | | | | | |
