## Supplementary material for "Effectiveness of the modified WHO labour care guide to detect prolonged and obstructed labour among women admitted at publicly funded facilities in rural Mbarara district, Southwestern Uganda: an ambispective cohort study": Table 1: Socio-demographic and obstetric characteristics of participant

| **Characteristics** | | **LCG (N=991)** | **Partograph (N=1,020)** | **p-value** |
| --- | --- | --- | --- | --- |
| **Health facility level** | | | | 0.12 |
|  | Health Centre IV | 845 (85.3%) | 894 (87.6%) |  |
|  | Health Centre III | 146 (14.7%) | 126 (12.4%) |  |
| **Maternal age in years (mean (SD)** | | 25.9 (5.8) | 25.9 (5.5) | 0.92 |
| **Age category** | | | | 0.47 |
|  | <20 | 90 (9.1%) | 77 (7.5%) |  |
|  | 20-24 | 310 (31.3%) | 306 (30.0%) |  |
|  | 25-34 | 317 (32.0%) | 334 (32.7%) |  |
|  | >34 | 274/991 (27.6%) | 303 (29.7%) |  |
| **Gravidity** | | | | 0.87 |
|  | 1 | 269 (27.1%) | 276 (27.1%) |  |
|  | 2-4 | 424 (42.8%) | 435 (42.6%) |  |
|  | >5 | 98 (9.9%) | 92 (9.0%) |  |
|  | Missing | 200 (20.2%) | 217 (21.3%) |  |
| **Parity** | | | | <0.001 |
|  | Primipara | 448 (45.2%) | 391 (38.3%) |  |
|  | Multipara (2-4) | 297 (30.0%) | 266 (26.1%) |  |
|  | Grand multipara (>5) | 27 (2.7%) | 33 (3.2%) |  |
|  | Missing para | 219 (22.1%) | 330 (32.4%) |  |
| **Mean gestational age at delivery (SD)** | | 39.3 (1.9) | 39.4 (1.7) | 0.39 |
| **Gestational age category (weeks)** | | | | 0.38 |
|  | 28-36 | 35 (3.5%) | 27 (2.6%) |  |
|  | 37-42 | 484 (48.8%) | 473 (46.4%) |  |
|  | >42 | 33 (3.3%) | 38 (3.7%) |  |
|  | Unknown | 439 (44.3%) | 482 (47.3%) |  |
| **HIV status** | | | | 0.11 |
|  | Negative | 814 (82.1%) | 804 (78.8%) |  |
|  | Positive | 70 (7.1%) | 75 (7.4%) |  |
|  | Unknown | 107 (10.8%) | 141 (13.8%) |  |
| **Mother was referred in** | | | | <0.001 |
|  | Yes | 54 (5.4%) | 9 (0.9%) |  |
|  | No | 937(94.6%) | 1,011(99.1%) |  |
| **Birth weight (Kg)** | | | | 0.42 |
|  | <2.5 | 28 (2.8%) | 27 (2.6%) |  |
|  | 2.5-<4 | 937 (94.6%) | 956 (93.7%) |  |
|  | ≥4 | 26 (2.6%) | 37 (3.6%) |  |
| **Baby sex** | | | | 0.15 |
|  | Male | 498/969 (51.4%) | 473/982 (48.2%) |  |
|  | Female | 471/969 (48.6%) | 509/982 (51.8%) |  |
| **Cadre of HCP conducting the delivery** | | | | <0.001 |
|  | Certificate Midwife | 621 (62.7%) | 794 (77.8%) |  |
|  | Diploma level Midwife/Nurse | 51 (5.1%) | 36 (3.6%) |  |
|  | Graduate Midwife/ or Medical Doctor | 204 (20.6%) | 61 (6%) |  |
|  | Others (students) | 115 (11.6%) | 129 (12.6) |  |
| *LCG: Labor care guide, HIV: Human Immunodeficiency Virus, SD: Standard Deviation, HCP: Health Care Provider* | | | | |
